# Factors associated with therapeutic inertia in individuals with type 2 diabetes mellitus started on basal insulin

**DOI:** 10.1101/2023.07.14.23292659

**Authors:** Sharmin Shabnam, Clare L Gillies, Melanie J Davies, Terry Dex, Eka Melson, Kamlesh Khunti, David R Webb, Francesco Zaccardi, Samuel Seidu

## Abstract

**Aim:** In this study we aim to identify the risk factors for treatment inertia in patients with type 2 diabetes mellitus (T2DM) who have been recently started on basal insulin (BI).

**Methods:** Using UK CPRD GOLD, we identified adults with T2DM with suboptimal glycaemia (HbA1c within 12 months of BI ≥7% (≥53 mmol/mol)). We used multivariable Cox regression model to describe the association between patient characteristics and the time to treatment intensification.

**Results:** A total of 12,556 patients were analysed. Compared to individuals aged <65 years, those aged ≥65 years had lower risk of treatment intensification (HR: 0.69; 95% CI: 0.64-0.73). Other risk factors included being female (0.93, 0.89-0.99), longer T2DM duration (0.99, 0.98-0.99), living in the most deprived areas (0.90, 0.83-0.98), being a current smoker (0.91, 0.84-0.98), having one (0.91, 0.85- 0.97) or more than one comorbidity (0.88, 0.82-0.94), and patients who were on metformin (0.71, 0.63-0.80), or 2nd generation sulphonylureas (0.85; 0.79-0.92) or DPP4 inhibitors (0.87, 0.82-0.93) compared to those who were not.

**Conclusion:** Therapeutic inertia still remains a major barrier, with multiple factors associated with delay in intensification. Interventions to overcome therapeutic inertia need to be implemented at both patient and health care professional level.

## Introduction

Type 2 diabetes mellitus (T2DM) is a global growing epidemic with significant impacts on individuals and healthcare systems. According to the International Diabetes Federation, 537 million people aged 20 to 79 years lived with diabetes mellitus in 2021, with an expected increase to 643 million by 2030 and 783 million by 2045, primarily in low- and middle-income countries.^1^ Individualised optimal glycated haemoglobin (HbA1c) target remains the cornerstone in the management of T2DM with evidence showing reductions in micro- and macrovascular complications following HbA1c lowering.^2–4^ Guidelines have recommended an individualised HbA1c target of <7% (<53 mmol/mol) for the majority of individuals with T2DM. HbA1c should then be monitored every 3-6 months and, if it remains above the target, treatment intensification with additional medications including insulin may be needed.^5–7^ Despite the recent advances in the treatment of T2DM, glycaemic control had been shown to be suboptimal globally.^8–10^ One key reason for this is therapeutic inertia, which is defined as ‘the failure of healthcare professionals to intensify therapy when the therapeutic goal (i.e., HbA1c) is not reached’.^11^

A meta-analysis including studies in both primary and secondary care showed that the median time to treatment intensification, following an above-the-target HbA1c measurement, was more than a year (range 0.3 to 7.2 years). This included patients who had their treatment intensified by addition of oral hypoglycaemics or insulin/other injectable therapies.^12^ Patients who are receiving injectable therapies seem to be disproportionately affected by therapeutic inertia. A retrospective cohort study in the UK showed that a quarter of patients with T2DM had insulin initiation delayed for at least 1.8 years, and half of the patients delayed starting insulin for almost 5 years after not achieving the target HbA1c range.^13^ These delays in treatment intensification represent significant risks to individuals with T2DM, as early and sustained glycaemic control had been shown to reduce long-term micro- and macrovascular complications.^14^ Therapeutic inertia in T2DM also represents a significant economic burden, with a study in Denmark showing a substantial healthcare cost of up to $50 million associated with delayed intensification.^15^ The IQVIA Core diabetes model in the UK also projected an increase in cost of £122 million and £930 million to complications and loss of productivity, associated with therapeutic inertia of 3 years.^16^

The reasons for therapeutic inertia are complex and can be categorised into healthcare professional-, patient-, and system-related factors.^17^ In terms of patient-related factors, these could be due to concerns about side effects, misunderstanding of treatment intensification, and presence of multimorbidity.^18^ In addition, the risk of falls and hypoglycaemia in patients on insulin further complicate treatment intensifications. However, despite recent advances in developing effective glucose-lowering therapies, the prevalence of therapeutic inertia is escalating, especially in patients on insulin.^13, 19^ In this regard, we conducted this study to identify the risk factors for treatment inertia in patients who have been recently started on basal insulin (BI).

## Methods

### Data Source and Study Population

This is a retrospective longitudinal cohort study using the Clinical Practice Research Datalink GOLD database, which routinely collects anonymized electronic health records from contributing primary care practices in the UK. The data used in this analysis is additionally linked to hospital admission records from the Hospital Episode Statistics (HES) Admitted Patient Care, mortality records from the Office for National Statistics (ONS) death registry, and patient level Index of Multiple Deprivation (IMD) 2015. Protocol for this study was approved by the Independent Scientific Advisory Committee (ISAC; protocol 19_047).

Patients were eligible if they had a first ever prescription of BI between the study period – 1 January 2004 and 31 December 2019 – and a clinical record of T2DM before the initiation of BI therapy. BI was defined as any long- or intermediate-acting or intravenous insulin. Patients were excluded if they had any record of type 1 diabetes in their clinical history. They were also excluded if they had any record of gestational or secondary diabetes before they started on BI. Patients were required to be aged 18 years or over, have at least 1 year of prior up-to-standard registration at the initiation of BI, and a valid HbA1c measurement within 12 months before and after BI initiation. Based on the last recorded value of HbA1c within 12 months following BI initiation (denoted as index date, Figure 1), patients were categorized into two groups – those with optimal glycaemic level (HbA1c <7% or <53 mmol/mol) and those with suboptimal glycaemic level (HbA1c ≥7% or ≥53 mmol/mol). For our analysis, we restricted the cohort to only those who have suboptimal glycaemic level. The end of follow-up was defined as either the date of the outcome, the date of death, end of ONS linkage date (31 May 2019), or the study end period (31 December 2019), whichever comes first; individuals were followed from the index date to the end of follow-up.

**Figure 1:**
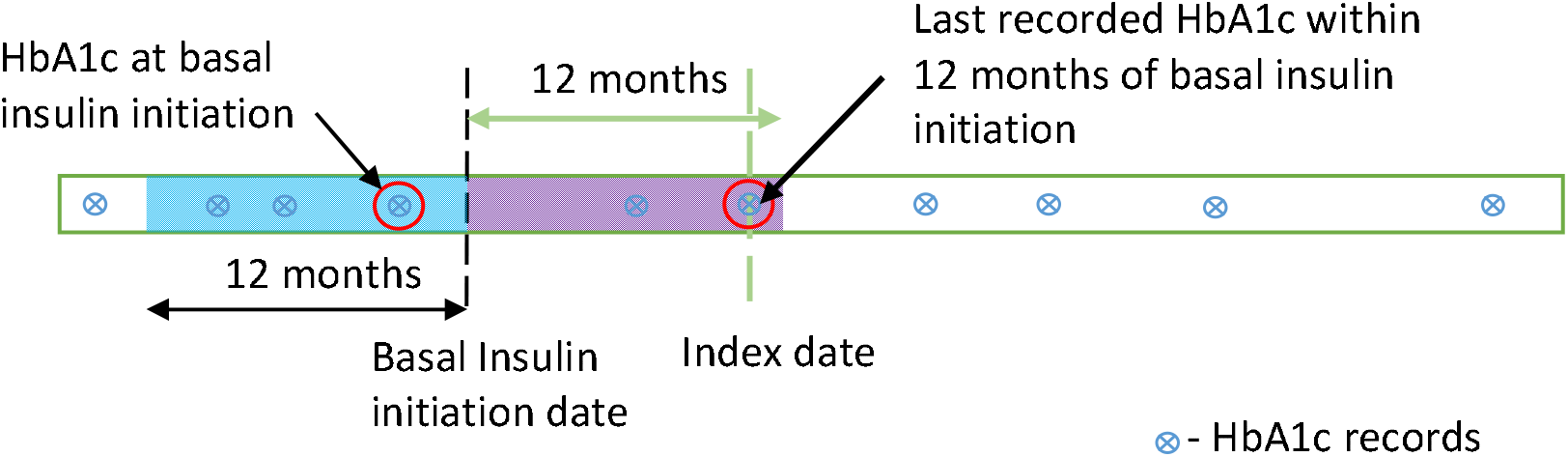
Definition of index date from the timeline of patient’s HbA1c record.

### Outcome

The study outcome is the intensification of treatment following suboptimal glycaemic level. The treatment intensification was defined as the occurrence of one of the following two events in patients’ medication records:

a. Addition of a short-acting insulin or changing to pre-mixed insulin, i.e., treatment was intensified when a patient had BI and started biphasic/prandial insulin or when a patient had biphasic insulin and then started prandial insulin.
b. Addition of other class of glucose-lowering drugs, such Oral Antidiabetic Drug (OAD) or Glucagon-Like Peptide 1 Receptor Agonists (GLP1-RA) while continuing with or without BI.

Prescription medications for each patient were coded in the CPRD records and were used to extract information regarding above mentioned insulin or glucose-lowering drugs. The time to treatment intensification was calculated from the index date to the earliest date of prescribing either (a) or (b) mentioned above. Patients in the no intensification group were censored at the end of their respective follow-up period.

### Predictor variables

As potential predictors of treatment intensification, we extracted the following information closest to the index date: demographic variables (age, sex, ethnicity, and deprivation), lifestyle (smoking status), clinical history (T2DM duration and prevalent comorbidities), clinical measurements (body mass index (BMI), HbA1c, Estimated Glomerular Filtration Rate (eGFR), blood pressure, total cholesterol) and history of glucose-lowering medications.

Age was calculated at the index date and ethnicity was primarily obtained from HES records. Missing or unknown ethnicities in HES were replaced with ethnicity records available in CPRD, and patients were categorised into four groups: White, South Asian, Black, and Mixed/Other. IMD quintiles were used to indicate patient deprivation level where a score of one represented the least deprived quintile and five the most deprived one. Smoking status was obtained from CPRD and categorised as current smoker, ex-smoker, and non-smoker. Systolic and diastolic blood pressure (mmHg), total cholesterol (mmol/l), HbA1c, and BMI (kg/m^2^) were ascertained from clinical measurements data in CPRD. Missing BMI values were calculated using same day recorded height and weight measurements; values of BMI <10 and >70 kg/m^2^ were excluded (considered clinically implausible) and patients were categorized into five groups: underweight (<18.5), normal weight (18.5 to 24.9), overweight (25 to 29.9), obesity (30 to 34.9), and severe obesity (≥35). Serum creatinine values in CPRD were used to calculate eGFR based on the CKD-EPI equation.^20^ For time-varying predictor variables, the closest record within 1 year before the index date was used.

T2DM duration is calculated from the date of the first recorded code of T2DM in CPRD to the index date. Prevalent comorbidities at the index date for each patient were obtained from both hospitalization and primary care records, including: diabetic retinopathy, diabetic neuropathy, hypoglycaemia, hyperglycaemia, diabetes with peripheral circulatory disorders, gangrene, lower extremity amputation, diabetes with unspecified complication, diabetes mellitus without mention of complications, ischemic heart disease, acute myocardial infarction, old myocardial infarction, acute coronary syndrome or unstable angina, any stroke, transient cerebral ischemia and related syndromes, congestive heart failure, peripheral vascular disease surgery or chronic renal insufficiency; and categorized into none, one, two or more comorbidities. From prescription records in CPRD, we also collected information on history of glucose-lowering medications at the index date, including: metformin, 2^nd^ generation sulphonylureas, GLP1-RA, thiazolidinediones, meglitinides, alpha-glucosidase inhibitors, sodium-glucose transport protein 2 (SGLT2) inhibitors, and dipeptidyl peptidase-4 (DPP4) inhibitors.

### Statistical Analysis

Descriptive summary statistics of patient characteristics at the index date were presented for the overall cohort and based on the occurrence of intensification (yes/no): characteristics were described by numbers and percentages (categorical variables) and mean and standard deviation or median and interquartile range (continuous variables).

Complete-case multivariable Cox regression models were used to describe the association between patient characteristics and time to treatment intensification. We regarded these characteristics as potential predictors: age, T2DM duration, sex, ethnicity, IMD score, smoking status, BMI, HbA1c, eGFR, blood pressure (systolic), total cholesterol, prevalent comorbidities, and history of glucose-lowering medications. All analyses were conducted by using Python 3.8 and R Statistical Package version 4.1.2.

## Results

Study population characteristics A total of 12,556 patients on BI with suboptimal glycaemic level were included in our study (Figure 2): among them, 6,455 (51.4%) did not receive any treatment intensification following BI initiation during their follow-up period. The characteristics of patients who have received intensification, compared to those who have not, are summarised in Table 1. Mean follow-up period was lower for those who did not receive any treatment intensification (mean ± standard deviation SD: 5.5 ± 3.8 vs. 7.7 ± 3.8 years) compared to those who received. Median time to intensification for the latter group was 333 days (interquartile range IQR: 91.0, 840.0). Those who received no treatment intensification were older compared to those with their treatment intensified (68.0 ± 12.0 vs. 62.1 ± 12.5 years). Within the total follow-up period, 2334 patients (40.4%) in the younger age group and 4121 patients (60.8%) in the older age group still remained without any intensification.

**Figure 2:**
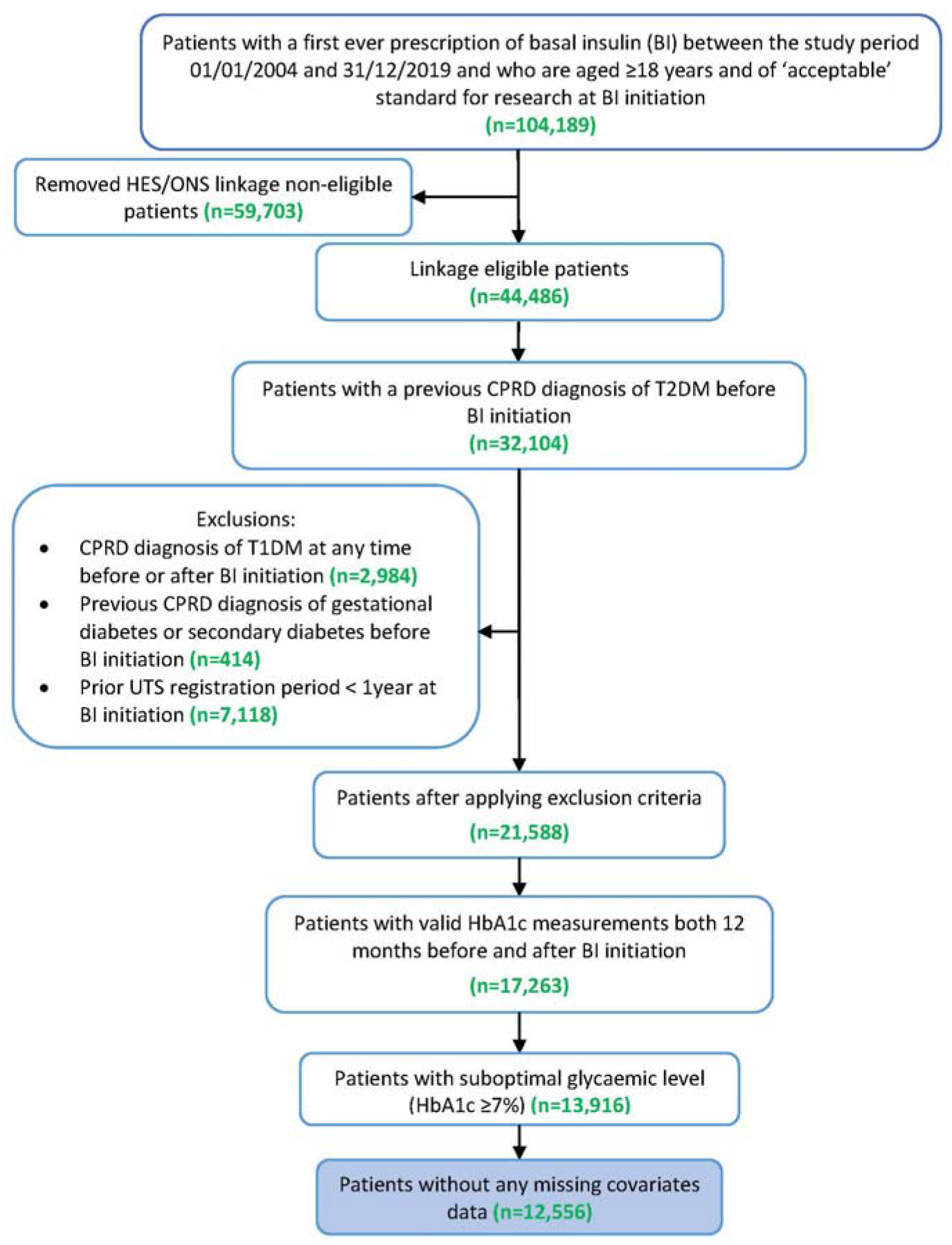
Study population flow diagram.

**Table 1:**
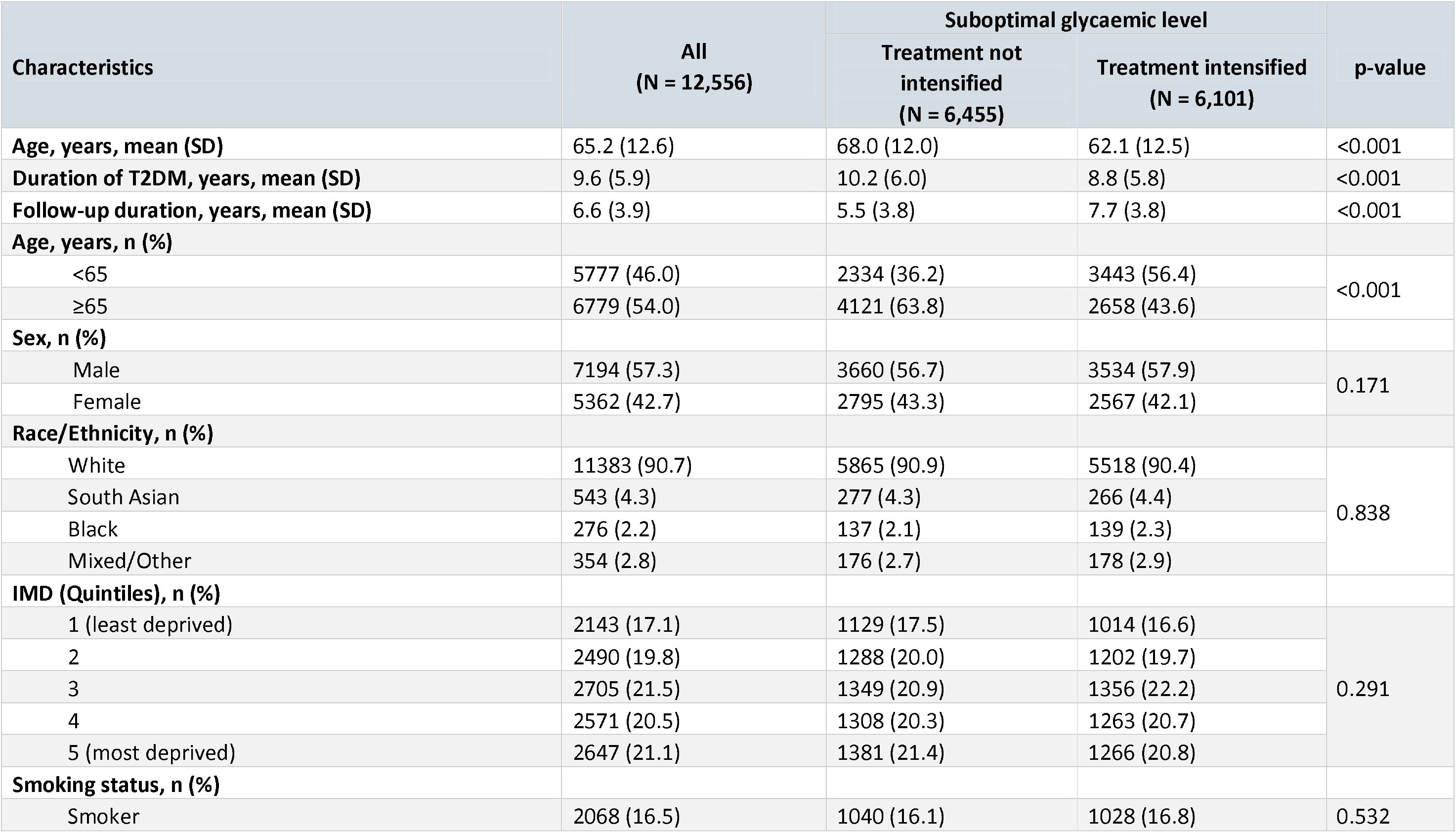
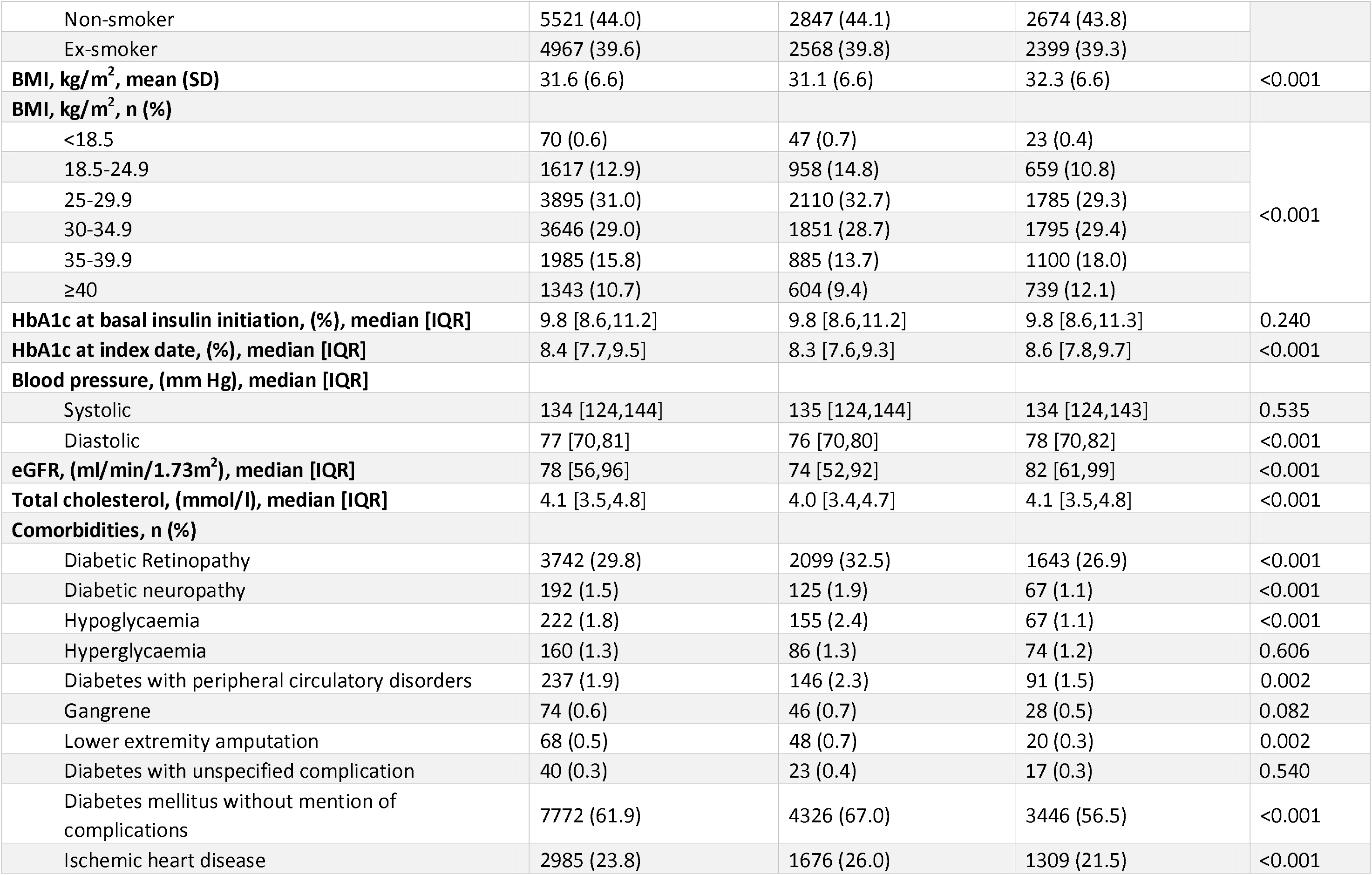
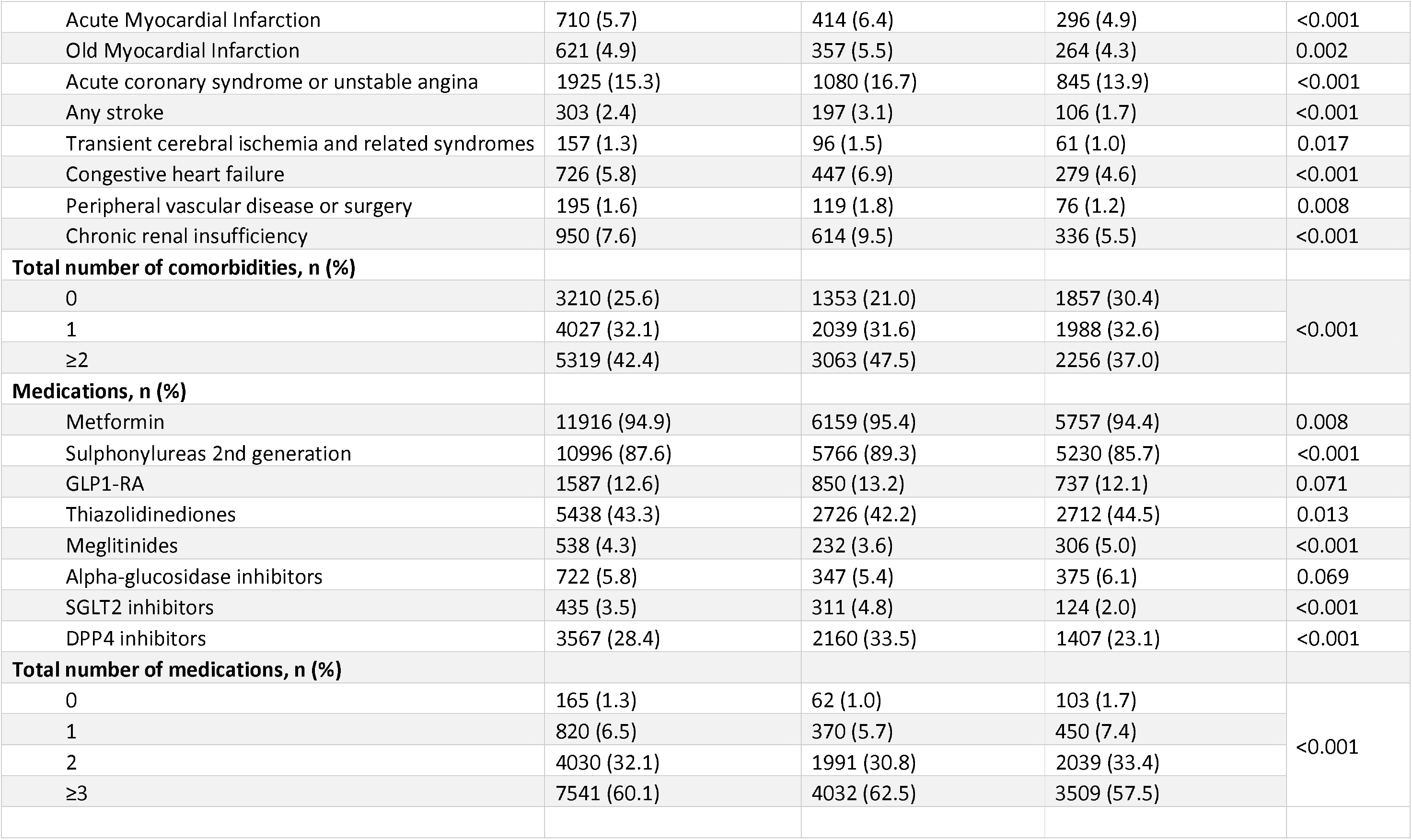
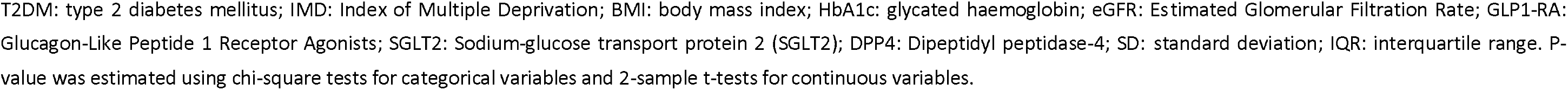
Characteristics of study cohort, overall and stratified by treatment groups.

| Characteristics | All<br>(N = 12,556) | Suboptimal glycaemic level |  | p-value |
| --- | --- | --- | --- | --- |
|  |  | Treatment not intensified<br>(N = 6,455) | Treatment intensified<br>(N = 6,101) |  |
| Age, years, mean (SD) | 65.2 (12.6) | 68.0 (12.0) | 62.1 (12.5) | <0.001 |
| Duration of T2DM, years, mean (SD) | 9.6 (5.9) | 10.2 (6.0) | 8.8 (5.8) | <0.001 |
| Follow-up duration, years, mean (SD) | 6.6 (3.9) | 5.5 (3.8) | 7.7 (3.8) | <0.001 |
| Age, years, n (%) |  |  |  |  |
| <65 | 5777 (46.0) | 2334 (36.2) | 3443 (56.4) | <0.001 |
| ≥65 | 6779 (54.0) | 4121 (63.8) | 2658 (43.6) |  |
| Sex, n (%) |  |  |  |  |
| Male | 7194 (57.3) | 3660 (56.7) | 3534 (57.9) | 0.171 |
| Female | 5362 (42.7) | 2795 (43.3) | 2567 (42.1) |  |
| Race/Ethnicity, n (%) |  |  |  |  |
| White | 11383 (90.7) | 5865 (90.9) | 5518 (90.4) | 0.838 |
| South Asian | 543 (4.3) | 277 (4.3) | 266 (4.4) |  |
| Black | 276 (2.2) | 137 (2.1) | 139 (2.3) |  |
| Mixed/Other | 354 (2.8) | 176 (2.7) | 178 (2.9) |  |
| IMD (Quintiles), n (%) |  |  |  |  |
| 1 (least deprived) | 2143 (17.1) | 1129 (17.5) | 1014 (16.6) | 0.291 |
| 2 | 2490 (19.8) | 1288 (20.0) | 1202 (19.7) |  |
| 3 | 2705 (21.5) | 1349 (20.9) | 1356 (22.2) |  |
| 4 | 2571 (20.5) | 1308 (20.3) | 1263 (20.7) |  |
| 5 (most deprived) | 2647 (21.1) | 1381 (21.4) | 1266 (20.8) |  |
| Smoking status, n (%) |  |  |  |  |
| Smoker | 2068 (16.5) | 1040 (16.1) | 1028 (16.8) | 0.532 |
| Non-smoker | 5521 (44.0) | 2847 (44.1) | 2674 (43.8) |  |
| Ex-smoker | 4967 (39.6) | 2568 (39.8) | 2399 (39.3) |  |
| **BMI, kg/m<sup>2</sup>, mean (SD)** | 31.6 (6.6) | 31.1 (6.6) | 32.3 (6.6) | <0.001 |
| **BMI, kg/m<sup>2</sup>, n (%)** |  |  |  |  |
| <18.5 | 70 (0.6) | 47 (0.7) | 23 (0.4) | <0.001 |
| 18.5-24.9 | 1617 (12.9) | 958 (14.8) | 659 (10.8) |  |
| 25-29.9 | 3895 (31.0) | 2110 (32.7) | 1785 (29.3) |  |
| 30-34.9 | 3646 (29.0) | 1851 (28.7) | 1795 (29.4) |  |
| 35-39.9 | 1985 (15.8) | 885 (13.7) | 1100 (18.0) |  |
| ≥40 | 1343 (10.7) | 604 (9.4) | 739 (12.1) |  |
| **HbA1c at basal insulin initiation, (%), median [IQR]** | 9.8 [8.6,11.2] | 9.8 [8.6,11.2] | 9.8 [8.6,11.3] | 0.240 |
| **HbA1c at index date, (%), median [IQR]** | 8.4 [7.7,9.5] | 8.3 [7.6,9.3] | 8.6 [7.8,9.7] | <0.001 |
| **Blood pressure, (mm Hg), median [IQR]** |  |  |  |  |
| Systolic | 134 [124,144] | 135 [124,144] | 134 [124,143] | 0.535 |
| Diastolic | 77 [70,81] | 76 [70,80] | 78 [70,82] | <0.001 |
| **eGFR, (ml/min/1.73m<sup>2</sup>), median [IQR]** | 78 [56,96] | 74 [52,92] | 82 [61,99] | <0.001 |
| **Total cholesterol, (mmol/l), median [IQR]** | 4.1 [3.5,4.8] | 4.0 [3.4,4.7] | 4.1 [3.5,4.8] | <0.001 |
| **Comorbidities, n (%)** |  |  |  |  |
| Diabetic Retinopathy | 3742 (29.8) | 2099 (32.5) | 1643 (26.9) | <0.001 |
| Diabetic neuropathy | 192 (1.5) | 125 (1.9) | 67 (1.1) | <0.001 |
| Hypoglycaemia | 222 (1.8) | 155 (2.4) | 67 (1.1) | <0.001 |
| Hyperglycaemia | 160 (1.3) | 86 (1.3) | 74 (1.2) | 0.606 |
| Diabetes with peripheral circulatory disorders | 237 (1.9) | 146 (2.3) | 91 (1.5) | 0.002 |
| Gangrene | 74 (0.6) | 46 (0.7) | 28 (0.5) | 0.082 |
| Lower extremity amputation | 68 (0.5) | 48 (0.7) | 20 (0.3) | 0.002 |
| Diabetes with unspecified complication | 40 (0.3) | 23 (0.4) | 17 (0.3) | 0.540 |
| Diabetes mellitus without mention of complications | 7772 (61.9) | 4326 (67.0) | 3446 (56.5) | <0.001 |
| Ischemic heart disease | 2985 (23.8) | 1676 (26.0) | 1309 (21.5) | <0.001 |
| Acute Myocardial Infarction | 710 (5.7) | 414 (6.4) | 296 (4.9) | <0.001 |
| Old Myocardial Infarction | 621 (4.9) | 357 (5.5) | 264 (4.3) | 0.002 |
| Acute coronary syndrome or unstable angina | 1925 (15.3) | 1080 (16.7) | 845 (13.9) | <0.001 |
| Any stroke | 303 (2.4) | 197 (3.1) | 106 (1.7) | <0.001 |
| Transient cerebral ischemia and related syndromes | 157 (1.3) | 96 (1.5) | 61 (1.0) | 0.017 |
| Congestive heart failure | 726 (5.8) | 447 (6.9) | 279 (4.6) | <0.001 |
| Peripheral vascular disease or surgery | 195 (1.6) | 119 (1.8) | 76 (1.2) | 0.008 |
| Chronic renal insufficiency | 950 (7.6) | 614 (9.5) | 336 (5.5) | <0.001 |
| **Total number of comorbidities, n (%)** |  |  |  |  |
| 0 | 3210 (25.6) | 1353 (21.0) | 1857 (30.4) | <0.001 |
| 1 | 4027 (32.1) | 2039 (31.6) | 1988 (32.6) |  |
| ≥2 | 5319 (42.4) | 3063 (47.5) | 2256 (37.0) |  |
| **Medications, n (%)** |  |  |  |  |
| Metformin | 11916 (94.9) | 6159 (95.4) | 5757 (94.4) | 0.008 |
| Sulphonylureas 2nd generation | 10996 (87.6) | 5766 (89.3) | 5230 (85.7) | <0.001 |
| GLP1-RA | 1587 (12.6) | 850 (13.2) | 737 (12.1) | 0.071 |
| Thiazolidinediones | 5438 (43.3) | 2726 (42.2) | 2712 (44.5) | 0.013 |
| Meglitinides | 538 (4.3) | 232 (3.6) | 306 (5.0) | <0.001 |
| Alpha-glucosidase inhibitors | 722 (5.8) | 347 (5.4) | 375 (6.1) | 0.069 |
| SGLT2 inhibitors | 435 (3.5) | 311 (4.8) | 124 (2.0) | <0.001 |
| DPP4 inhibitors | 3567 (28.4) | 2160 (33.5) | 1407 (23.1) | <0.001 |
| **Total number of medications, n (%)** |  |  |  |  |
| 0 | 165 (1.3) | 62 (1.0) | 103 (1.7) | <0.001 |
| 1 | 820 (6.5) | 370 (5.7) | 450 (7.4) |  |
| 2 | 4030 (32.1) | 1991 (30.8) | 2039 (33.4) |  |
| ≥3 | 7541 (60.1) | 4032 (62.5) | 3509 (57.5) |  |

T2DM: type 2 diabetes mellitus; IMD: Index of Multiple Deprivation; BMI: body mass index; HbA1c: glycated haemoglobin; eGFR: Estimated Glomerular Filtration Rate; GLP1-RA: Glucagon-Like Peptide 1 Receptor Agonists; SGLT2: Sodium-glucose transport protein 2 (SGLT2); DPP4: Dipeptidyl peptidase-4; SD: standard deviation; IQR: interquartile range. P-value was estimated using chi-square tests for categorical variables and 2-sample t-tests for continuous variables.

Figures 3 and 4 show the Kaplan-Meier curves for the probability of intensification, overall and by age groups. Overall, intensification occurred till about 15 years when the probability of intensification reached a maximum of 58.0% (Figure 3). Additionally, those under the age of 65 were relatively more likely to receive intensification much earlier, with higher proportions of them intensified at any follow-up time compared to those aged 65 years or more (Figure 4). At 4 years of follow-up after BI initiation, in those aged ≥65 years, the probability of intensification was 38.0%, after which the probability of them receiving any further intensification was rather stable, with only reaching to 50.2% in the following 10 years. Conversely, for those aged <65 years, at 4 years, the probability of intensification was 54.7% which increased to only 66.2% at the end of follow-up. (Figure 4).

**Figure 3:**
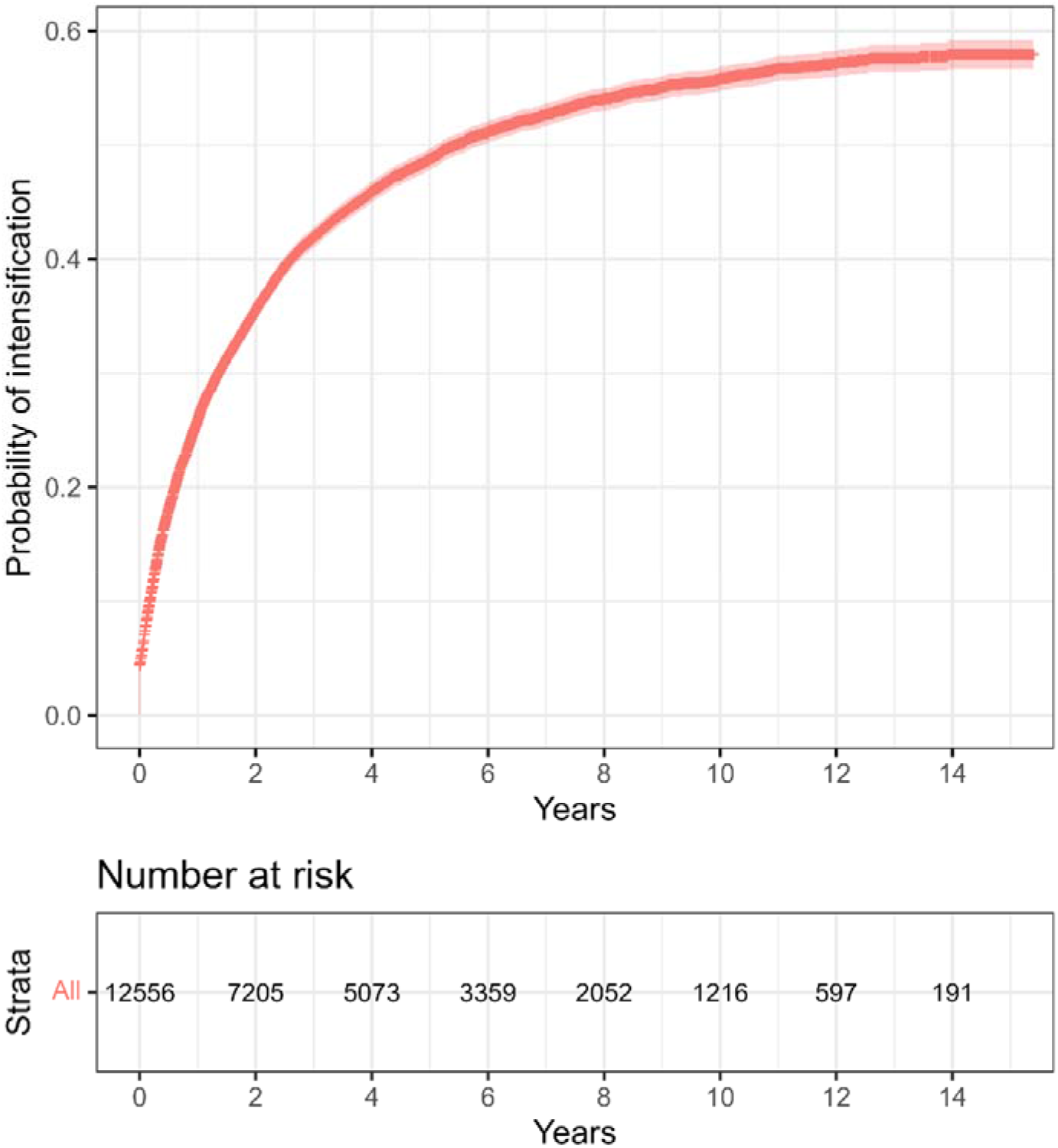
Kaplan–Meier probability of intensification.

**Figure 4:**
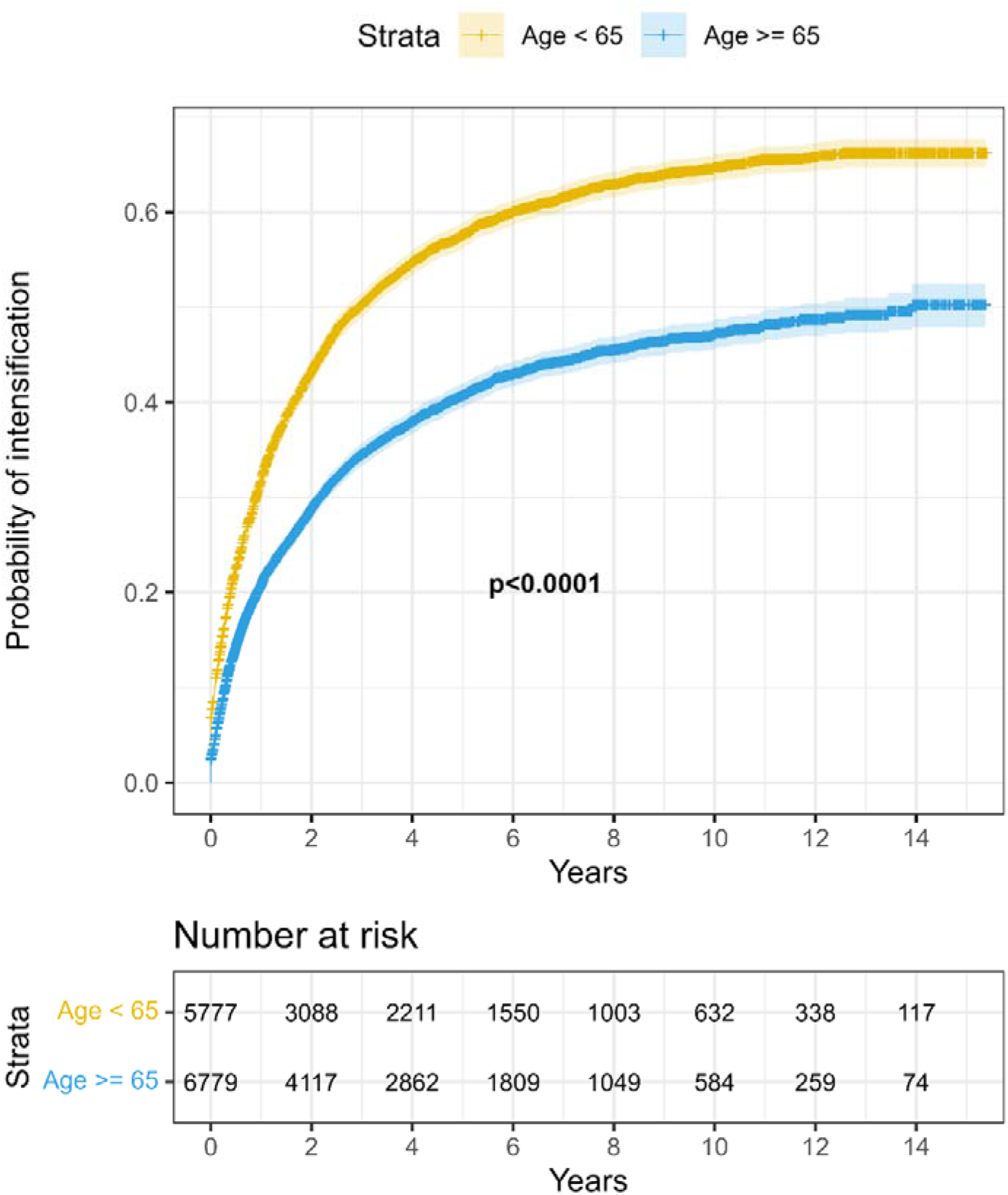
Kaplan–Meier probability of intensification by age groups.

Patients who were not intensified had a longer duration of T2DM (10.2 ± 6.0 years vs. 8.8 ± 5.8 years), and lower BMI (31.1 ± 6.6 years vs. 32.3 ± 6.6 years) compared to those who were intensified. Regarding prevalent co-morbidities, there were higher proportions of patients who were not intensified compared to those intensified for the following conditions: diabetic retinopathy (32.5% vs 26.9%), diabetic neuropathy (1.9% vs. 1.1%), hypoglycaemia (2.4% vs. 1.1%), ischaemic heart diseases (26.0% vs. 21.5%), diabetes mellitus without mention of complications (67.0% vs. 56.5%), acute myocardial infarction (6.4% vs. 4.9%), acute coronary syndrome or unstable angina (16.7 vs. 3.9%), stroke (3.1% vs. 1.7%), congestive heart failure (6.9% vs. 4.6%), and chronic renal insufficiency (9.5% vs. 5.5%). Patients who had two or more co-morbidities represented a higher proportion of those who were not intensified vs those intensified (47.5% vs 37.0%).

Furthermore, a higher proportion of patients were on sulphonylureas (89.3% vs 85.7%), SGLT2 inhibitors (4.8% vs 2.0%), and DPP4 inhibitors (33.5% vs 23.1%) among those who were not intensified, compared to those intensified. Those on 3 or more medications had a higher proportion of patients who were not intensified (62.5% vs. 57.5%) compared to those intensified.

### Risk factors associated with time to treatment intensification

In a multivariable Cox model, patients with age ≥65 years were at a higher risk of a longer time to intensification compared to those younger than 65 years (adjusted Hazard Ratio (HR): 0.69, 95% Confidence Intervals (CI): 0.64-0.73; Table 2). Other risk factors included being female (compared to male; 0.93, 0.89-0.99), longer T2DM duration (for each additional year of diabetes: 0.99, 0.98-0.99), living in the most deprived areas (compared to living in the least; 0.90, 0.83-0.98), being a current smoker (compared to those who do not smoke; 0.91, 0.84-0.98), having one (compared to those who do not have: 0.91, 0.85-0.97) or more than one comorbidities (0.88, 0.82-0.94), and patients who were on metformin (0.71, 0.63-0.80), or 2nd generation sulphonylureas (0.85; 0.79-0.92) or DPP4 inhibitors (0.87, 0.82-0.93) compared to those who were not. Higher HbA1c (per 1 unit change of HbA1c in %: 1.09, 1.08-1.11), total cholesterol (per 1 unit change of cholesterol in mmol/l: 1.02, 1.00-1.04), BMI (compared to those with normal weight – those with obesity (30 to 34.9 kg/m2): 1.12, 1.02-1.23, and those with severe obesity (≥35 kg/m2): 1.24, 1.13-1.37), and prescription of meglitinides (1.24, 1.11-1.39) appeared to be associated with a shorter time to treatment intensification.

**Table 2:** Association of risk factors with time to treatment intensification.

| Characteristics | Adjusted HR (95% CI) |
| --- | --- |
| **Age** |  |
| < 65 years | Reference |
| ≥ 65 years | **0.69 (0.64, 0.73)** |
| **T2DM duration (per 1-year increase)** | **0.99 (0.98, 0.99)** |
| **Sex** |  |
| Male | Reference |
| Female | **0.93 (0.89, 0.99)** |
| **Race/Ethnicity** |  |
| White | Reference |
| Black | 0.96 (0.81, 1.13) |
| Mixed/Other | 0.95 (0.82, 1.11) |
| South Asian | 0.91 (0.80, 1.03) |
| **IMD (Quintiles)** |  |
| 1 (least deprived) | Reference |
| 2 | 0.99 (0.91, 1.08) |
| 3 | 1.00 (0.92, 1.08) |
| 4 | 0.94 (0.87, 1.02) |
| 5 (most deprived) | **0.90 (0.83, 0.98)** |
| **Smoking status** |  |
| Non-smoker | Reference |
| Ex-smoker | 1.03 (0.97, 1.09) |
| Smoker | **0.91 (0.84, 0.98)** |
| **BMI** |  |
| Normal weight (18.5 to 24.9 kg/m<sup>2</sup>) | Reference |
| Underweight (<18.5 kg/m<sup>2</sup>) | 0.89 (0.59, 1.35) |
| Overweight (25 to 29.9 kg/m<sup>2</sup>) | 1.07 (0.98, 1.17) |
| Obesity (30 to 34.9 kg/m<sup>2</sup>) | **1.12 (1.02, 1.23)** |
| Severe Obesity (≥35 kg/m<sup>2</sup>) | **1.24 (1.13, 1.37)** |
| **HbA1c (per 1 unit change of HbA1c in %)** | **1.09 (1.08, 1.11)** |
| **Blood pressure (per 1 unit change of blood pressure in mmHg)** |  |
| Systolic | 1.00 (1.00, 1.00) |
| **eGFR (per 1 unit change of eGFR in ml/min/1.73m<sup>2</sup>)** | 1.00 (1.00, 1.00) |
| **Total cholesterol (per 1 unit change of total cholesterol in mmol/l)** | **1.02 (1.00, 1.04)** |
| **Comorbidities** |  |
| None | Reference |
| One | **0.91 (0.85, 0.97)** |
| Two or more | **0.88 (0.82, 0.94)** |
| **Medications (yes vs no)** |  |
| Metformin | **0.71 (0.63, 0.80)** |
| Sulphonylureas 2nd generation | **0.85 (0.79, 0.92)** |
| GLP1-RA | 0.93 (0.85, 1.01) |
| Thiazolidinediones | 0.99 (0.94, 1.04) |
| Meglitinides | **1.24 (1.11, 1.39)** |
| Alpha-glucosidase inhibitors | 1.08 (0.97, 1.21) |
| SGLT2 inhibitors | 0.86 (0.71, 1.03) |
| DPP4 inhibitors | **0.87 (0.82, 0.93)** |

T2DM: type 2 diabetes mellitus; IMD: Index of Multiple Deprivation; BMI: body mass index; HbA1c: glycated haemoglobin; eGFR: Estimated Glomerular Filtration Rate; GLP1-RA: Glucagon-Like Peptide 1 Receptor Agonists; SGLT2: Sodium-glucose transport protein 2 (SGLT2); DPP4: Dipeptidyl peptidase-4; HR: Hazard Ratio; CI: Confidence Interval.

## Discussion

This study shows that therapeutic inertia remained an overall issue in management of diabetes in patients who were receiving BI and have not achieved optimal glycaemic level within a year. Furthermore, older age, female sex, longer diabetes duration, socioeconomic deprivation, being a current smoker, comorbidity, and being on metformin or 2nd generation sulphonylureas or DPP4 inhibitors were each associated with failure of treatment intensification following BI initiation. We have also found that, while those who were older (≥65 years) had a significantly lower likelihood of treatment intensification than those who were younger, a considerable proportion of people (about 4 in 10) in the younger group also did not receive any intensification following suboptimal glycaemia during the entire follow-up period. Our study highlights the therapeutic inertia and its key factors despite international guidelines recommending early intensification with glucose-lowering therapies, particularly in younger patients with less morbidity burden.

Several retrospective analyses have shown that patients with an age of ≥ 65 years being on 3 or more oral glucose-lowering drugs or BI were less likely to have their treatment intensified.^21, 22^ Having more co-morbidities was also associated with delay in treatment intensification.^23^ A large UK retrospective cohort study involving 80,000 patients with T2DM demonstrated that most patients remain with poor glycaemic control for several years before treatment intensification.^12^ Delays in the treatment in this cohort of patients could possibly be attributed to reasons such as to prevent polypharmacy and the potential risk of hypoglycaemia. A retrospective study by Khunti et al.^19^ in patients who have recently been started on BI showed a median of 4.3 years to treatment intensification; this delay was also associated with increasing age, numbers of oral anti glycaemic used, co-morbidities, and diabetes duration. A smaller study later showed a better time to treatment intensification of 58 days.^24^ However, most of the patients remained outside their target range following intensification. The overall evidence would therefore suggest that there is a well-defined group of patients with certain characteristics that were at a higher risk of treatment inertia and our study, in this regard, reinforces that some of the risk is driven by patient-level factors.

In another study, McCoy et al. noted that with increasing age and the build-up of diabetes- concordant and, to a lesser extent, discordant, and progressed comorbid conditions, the proportion of individuals achieving low HbA1c levels, and the odds of doing so using insulin, increased.^25^ Such increased likelihood of achieving low HbA1c level could explain the lower probability of intensification noted in our study. Conversely McCoy et al. noted that people who were younger and had fewer comorbidities were less likely to achieve low glycaemic level, and this could explain why we observed a relatively higher rate of intensification in those under 65 in our study.^25^

The risk of an emergency department visits or hospitalisation for hypoglycaemia had been noted to be the greatest in patients with diabetes, multiple comorbidities, prior severe hypoglycaemia, and sulphonylureas and/or insulin use.^26^ Three quarters of healthcare professionals in a survey reported hypoglycaemia as the most common barrier to insulin therapy.^27^ This could explain the cautious management approach in older patients in our study. However, the risks of hypoglycaemia in several studies involving intensification/optimisation of insulin therapy are low, ranging from 0-1.7%. The risks of hypoglycaemia could be further reduced by appropriate glucose-lowering review with discontinuation of insulin secretagogues, such as sulphonylureas or meglitinides as recommended by guidelines.^6, 28^ Our study also showed that those who were younger and not intensified, were prescribed sulphonylureas, which could possibly contribute to therapeutic inertia due to reluctance to transition into other form of medication.

With regards to the key demographic groups who were still experiencing treatment inertia in our study, interventions would need be personalized, appropriate, and multifactorial to improve the quality of T2DM care. Several studies have attempted to suggest ways to overcome therapeutic inertia.^17, 29^ Apart from patient-level factors, healthcare professional-related factors also remain a barrier to treatment intensification, especially with the use of insulin. This was shown in the ‘Management of Diabetes in Future Years’ (MODIFY) survey, revealing that 30% of healthcare professionals in primary care have no or very little experience in intensifying insulin therapy.^30^ Healthcare professional factors represent many of the causes of treatment inertia.^17, 31^ In addition to lack of clinical experience, time constraints, competing demands, lack of knowledge, fear of side effects, and guideline variance are the most cited barriers in treatment intensification.^17^ The American Diabetes Association Standards of Care recommended consideration of a combination of injectables in patients who do not reach the target HbA1c.^5, 31^ This will either be by introducing rapid-acting insulin with the largest meal, changing the regimen to pre-mixed insulin or adding GLP1-RA. The decision to choose from adding another insulin or GLP1-RA will need to be individualised depending on rates of hypoglycaemia, cost, and patients’ BMI. It is important to note that GLP1-RA is only recommended by the National Institute for Health and Care Excellence (NICE) in the UK in patients with BMI of ≥35 kg/m^2^ with co-morbidities or those with BMI <35 kg/m^2^ for whom insulin is not an option.^6, 7^ These rules do not exist in the US and are not included in the American Diabetes Association Standards of Care. Regardless of the availability of GLP1-RA, optimisation of the use of insulin and its intensification remains essential to ensure optimal glycaemic level and possibly reduce micro- and macrovascular complications. Intensification following BI initiation with the stepwise and multiple daily injection approaches had been discussed thoroughly in a review by Abrahamson and Peters.^32^ Interventions to improve the knowledge of healthcare professionals with regards to (de-) intensification are needed in patients who have been recently started on BI.

There were a few limitations to this study. Firstly, the insulin doses were not known, and some patients might still be in the process of up titration depending on their fasting blood glucose when the HbA1c was taken. In the older population with frailty, appropriate treatment options may sometimes include de-intensification of treatment. In this study, our focus was on failure to intensify treatment and not de-intensification. We were also unable to get data on adherence to therapies. The strength of this study is utilizing a diverse and large sample size that is representative of the national population of the UK. This is especially important in individuals with T2DM, due to its cultural and social determinants, particularly around the use of insulin and other injectable therapies. This study also analysed a broad range of patient characteristics to identify potential risk factors.

## Conclusion

In conclusion, the present study showed that therapeutic inertia still remains a major barrier in patients with T2DM who started on BI. Despite the evidence of legacy benefits in glycaemic control, treatment inertia persisted in both younger and older individuals. Although those who were older had less likelihood of receiving intensification than those younger, about 4 in 10 people under the age of 65 also remained without any treatment intensification after initiation of BI. We have also identified some of the key patient-related factors for treatment inertia. It is therefore important to consider and remove the barriers to a timely insulin intensification, by recognising the characteristics of patients who seems most at risk of therapeutic inertia.

## Acknowledgments

Sh.S., C.L.G., F.Z., Sam.S., M.J.D., K.K. acknowledge the National Institute for Health Research Applied Research Collaborations – East Midlands (NIHR ARC – EM) and the NIHR Leicester Biomedical Research Centre. The views expressed in this publication are those of the author(s) and not necessarily those of the National Health Service (NHS), the NIHR or the Department of Health.

## Data sharing statement

This study was conducted using CPRD GOLD and linked data subject to protocol approval (ISAC Protocol 19_047). The data controller for CPRD (Department of Health and Social Care) does not allow sharing of raw data. Codes used to define the cohort, medical conditions, and medications are available at GitHub link: https://github.com/LRWE/factors_therapeutic_inertia_t2dm_basal_insulin.

